# Neuromuscular Electrical Stimulation Enhances Cerebral Oxygenation in Subacute Stroke: Insights Using functional Near Infrared Spectroscopy from the RETRAIN Phase 1 Study

**DOI:** 10.1101/2025.05.10.25327353

**Authors:** Kausik Chatterjee, Sandra Leason, Allam Harfoush, Yashika Arora, Anirban Dutta

**Affiliations:** Countess of Chester Hospital NHS Foundation Trust, UK; All India Institute of Medical Sciences Delhi, India; University of Birmingham, UK

**Author notes:** **Corresponding Author:** Anirban Dutta, PhD, Department of Metabolism and Systems Science, School of Medical Sciences, College of Medicine and Health, University of Birmingham, Birmingham B15 2TT, UK.

**Keywords:** stroke rehabilitation, neuromuscular electrical stimulation, cerebral perfusion, functional near-infrared spectroscopy, cerebral autoregulation

## Abstract

**Background:** Stroke is a leading cause of long-term disability worldwide. Neuromuscular electrical stimulation (NMES), such as the geko™ device, may enhance cerebral perfusion post-stroke by improving venous return. This study evaluated the cortical haemodynamic effects of NMES in subacute stroke survivors using functional near-infrared spectroscopy (fNIRS).

**Methods:** A prospective observational study was conducted in 18 patients (>7 days post-ischaemic stroke) receiving bilateral lower limb NMES. fNIRS measured changes in oxyhaemoglobin (HbO) and deoxyhaemoglobin (HbR) concentrations across varying NMES intensities and postures (supine, semi-supine, and upright). Data were analysed using a general linear model, with β-values reflecting haemodynamic response magnitude.

**Results:** NMES evoked significant cortical haemodynamic responses, with increased HbO observed across multiple sensorimotor regions. Upright posture significantly enhanced cortical tissue oxygenation (p=0.010). Higher stimulation intensities produced greater HbO responses, indicating a dose-dependent effect. Larger infarct size (>5 cm) was associated with increased haemodynamic response. These findings suggest NMES may influence neurovascular coupling and cerebral autoregulation during stroke recovery.

**Conclusions:** NMES via the geko™ device enhances cortical oxygenation in subacute stroke, particularly in upright positions and at higher intensities. The results support the potential use of NMES not only for venous thromboembolism prevention but also as an adjunctive strategy to promote cerebral perfusion and facilitate rehabilitation. Further trials are warranted to explore clinical efficacy and functional outcomes.

## Introduction

Stroke is a leading cause of mortality and long-term disability worldwide, with an estimated 15 million people suffering a stroke annually. Over 12 million people worldwide will experience their first stroke in a year, with 6.5 million deaths resulting from stroke-related causes^1^. In the UK alone, approximately 110,000 individuals experience a stroke each year, contributing to over £ 26 billion in total healthcare costs encompassing NHS and Personal Social Services (PSS) expenses, unpaid care, and productivity loss^2^. Despite advances in acute stroke care, a significant number of patients do not receive timely reperfusion therapies such as thrombolysis or thrombectomy due to narrow treatment windows and eligibility restrictions^3^. Moreover, even among those who achieve successful recanalization, outcomes can remain poor due to factors such as impaired collateral circulation and compromised cerebral autoregulation^4^.

The pathophysiology of cerebral injury following ischemic stroke is complex and time dependent. Cerebral function relies on continuous blood flow to deliver oxygen and glucose. Interruption of cerebral blood flow (CBF), even for a few minutes, can lead to irreversible tissue damage. The body’s intrinsic protective mechanism, cerebral autoregulation (CA), attempts to maintain constant CBF across a range of blood pressures by altering vascular tone ^5^. However, ischemic stroke disrupts this balance. During the acute phase, the brain undergoes a series of compensatory phases—from autoregulatory vasodilation to increased oxygen extraction fraction (OEF), and finally metabolic failure in the ischemic core^6^. The penumbral region, where tissue is functionally impaired but potentially salvageable, becomes the main target of reperfusion therapy^7^.

Monitoring and enhancing cerebral perfusion in stroke patients are of critical importance, particularly in the subacute phase where hemodynamic factors can still influence outcomes. Dynamic cerebral autoregulation (dCA)—the brain’s ability to adjust blood vessel diameter in real-time to stabilize CBF—is increasingly recognized as a crucial physiological process during stroke recovery^8^. While traditionally assessed using transcranial Doppler (TCD), functional near-infrared spectroscopy (fNIRS) has emerged as a promising non-invasive alternative^9^. fNIRS measures cortical hemodynamics through changes in oxy- and deoxyhemoglobin concentrations, enabling real-time assessment of blood volume and oxygenation in superficial cortical regions^10^. Compared to TCD, fNIRS is easier to administer and better tolerated, although it is limited to cortical surface regions.

Given the limitations of conventional approaches to augment CBF, there is growing interest in novel methods such as neuromuscular electrical stimulation (NMES)^11,12^. Lower-limb NMES may enhance cerebral perfusion by increasing venous return and cardiac preload, thereby elevating cardiac output and internal carotid artery (ICA) blood flow^12^. A 2021 Japanese study^13^ found that calf and thigh NMES increased ICA blood flow by approximately 12%, while vertebral artery flow remained unchanged. This effect was strongly associated with a rise in end-tidal CO_2_ (R = 0.74), suggesting that NMES-induced hypercapnia led to cerebral vasodilation, particularly in the anterior circulation, due to the passive nature of the exercise. Here, the geko™ Electro-Stimulation Device (Firstkind Ltd, UK), originally developed for venous thromboembolism (VTE) prevention, delivers low-voltage electrical impulses to the common peroneal nerve, reflex activating the calf muscle pump and potentially enhancing venous return and cerebral perfusion^14^. Previous studies in healthy volunteers have shown that the geko™ device significantly increases cerebral blood volume^11^; however, its effects in subacute stroke remained unverified.

The RETRAIN trial (Restoring Effective Tissue perfusion using Neuromuscular Stimulation) was conceived to address this gap. The Phase 1 study (NCT06614400) investigates the immediate hemodynamic effects of varying intensities of NMES on cortical haemodynamics in subacute stroke patients using fNIRS. The primary objective is to identify the optimal NMES intensity and posture for maximizing cortical tissue oxygenation. Phase 2 of the trial will compare NMES to intermittent pneumatic compression (IPC)—another VTE prevention method—in the hyperacute stroke phase (<48 hours), further evaluating their relative efficacy and safety.

Ultimately, RETRAIN aims to establish whether common peroneal NMES can serve as a viable therapeutic adjunct to enhance cerebral perfusion and improve functional outcomes in stroke survivors. This paper presents primary outcome from the Phase 1 trial, offering novel insights into the physiological responses to NMES and informing future strategies for subacute stroke rehabilitation.

## Methodology

### Study Design

The RETRAIN Phase 1 trial (NCT06614400) was a prospective, observational study conducted at a single centre: the Stroke Rehabilitation Unit at Ellesmere Port Hospital, Chester, UK. This study (trial flow chart shown in Figure 1) was designed following Good Clinical Practice (GCP) guidelines and the Declaration of Helsinki, with appropriate ethical approvals secured prior to recruitment. Written informed consent was obtained from all participants.

**Figure 1:**
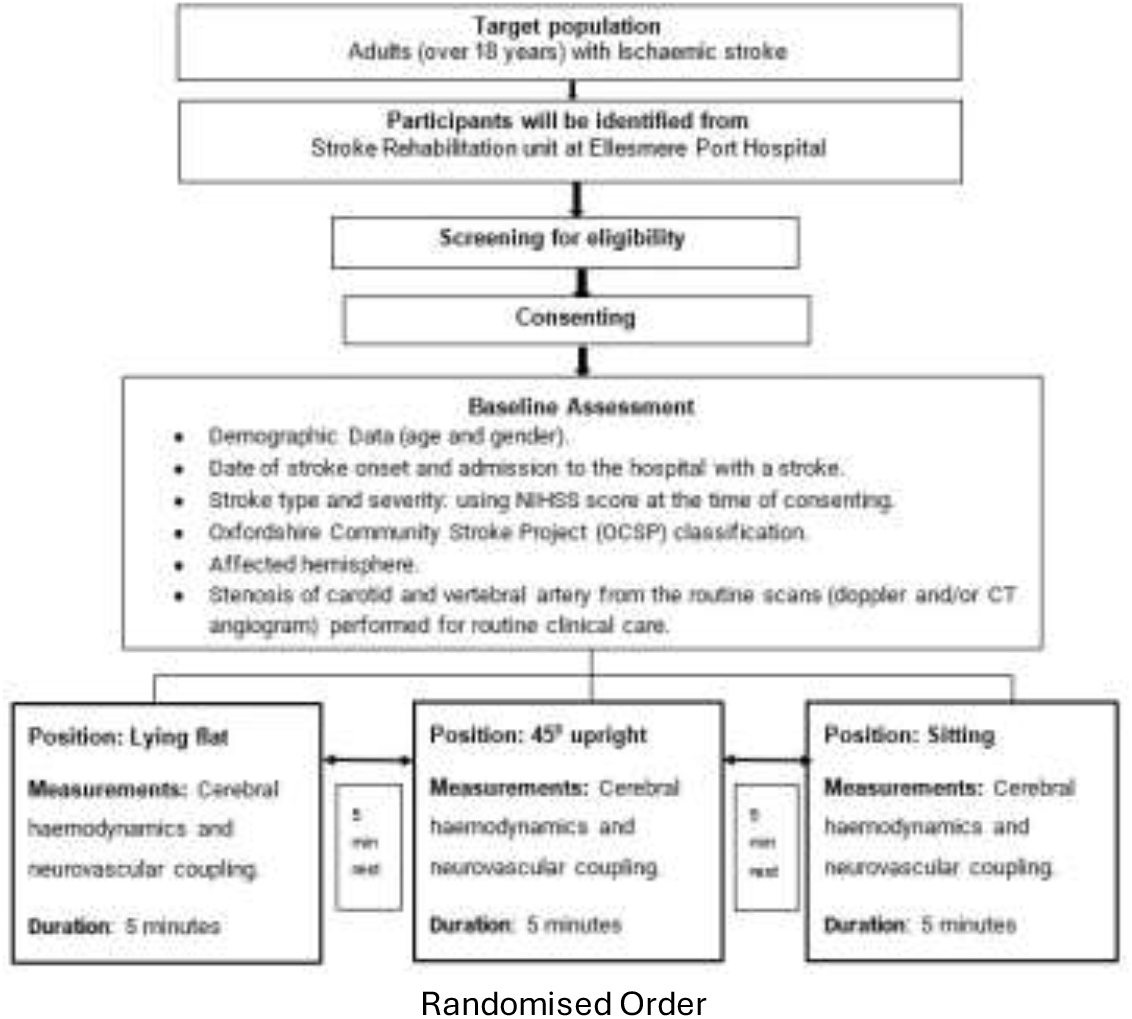
RETRAIN trial flow chart Phase 1

### Participants

Participants comprised adults over 18 years of age who had suffered a confirmed ischaemic stroke (validated via computed tomography [CT] or magnetic resonance imaging [MRI]) and were more than seven days post-stroke onset. Patients eligible for enrolment needed to be capable of safely assuming three specific postural positions (supine, semi-supine at 45 degrees, and seated upright) with assistance, and already receiving NMES via the geko™ device as part of routine clinical management for VTE prevention.

Exclusion criteria were comprehensive, including individuals with recent strokes within seven days, transient ischaemic attacks (TIA), epilepsy, peripheral neuropathy, recent lower limb amputations, concurrent use of other neuromodulatory devices, or an inability to provide informed consent due to cognitive impairment or communication difficulties.

### Procedures

Participants underwent thorough baseline assessments upon recruitment, including detailed demographic data (age, gender), stroke severity classification utilising the National Institutes of Health Stroke Scale (NIHSS), and relevant clinical vascular imaging findings obtained through routine care (such as Doppler ultrasound or CT angiography).

Eligible patients were identified by a qualified stroke physician. Once positioned securely, bilateral functional near-infrared spectroscopy (fNIRS) probes from the NIRSport2 fNIRS system (NIRx Medical Technologies, LLC, Glen Head, NY, USA) were placed on the sensorimotor area of the head (see Figure 2 for montage). Careful positioning avoided hair interference and sinus regions to optimize signal quality. Probes were secured using double-sided adhesive tape and elastic bandages for stable contact throughout the measurements.

**Figure 2:**
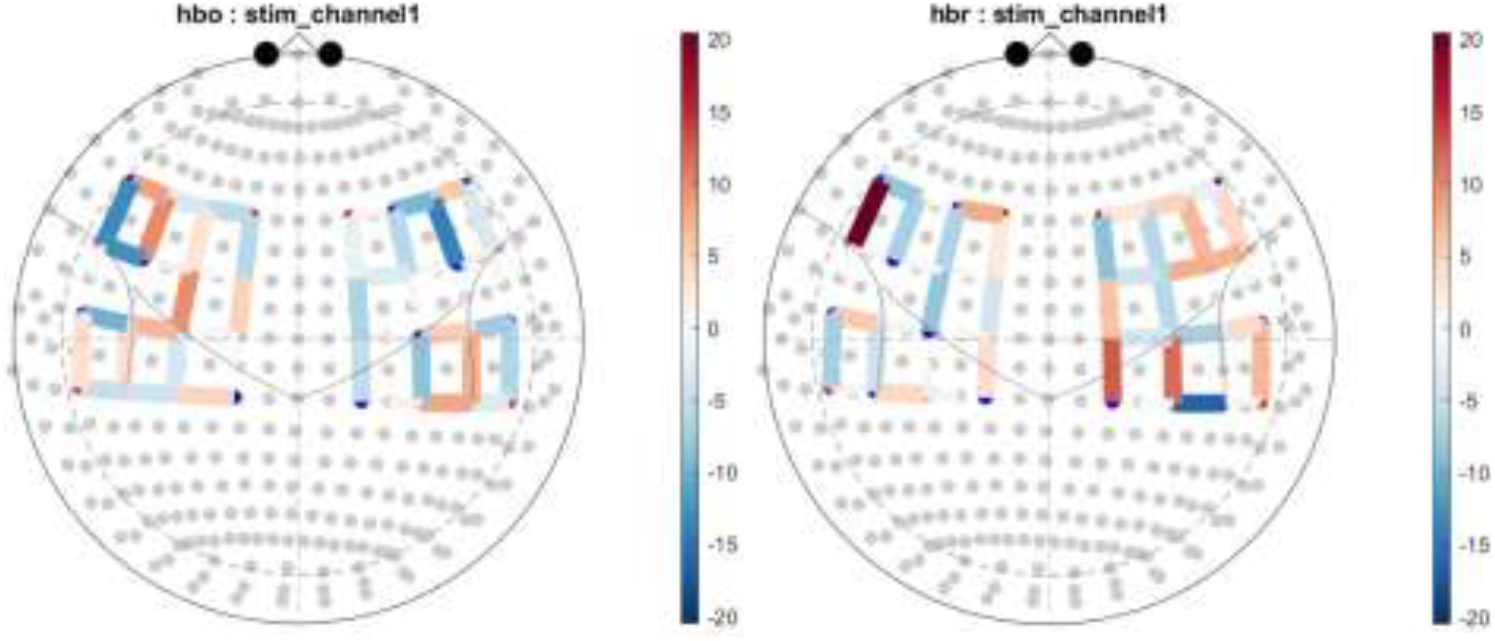
Topographical statistical parametric maps showing NMES-evoked changes in oxyhemoglobin (hbo) and deoxyhemoglobin (hbr) concentrations from pre-stimulation baseline across the cortex with significance determined by a false discovery rate (q<0.05).

The experimental intervention involved transcutaneous neuromuscular electrical stimulation (NMES) using the geko™ T-3 device (Firstkind Ltd, UK) bilaterally applied to the lower limbs to stimulate the common peroneal nerve. Each participant underwent stimulation across five distinct intensity levels: optimal (individualized standard clinical setting inducing dorsiflexion), one intensity level above optimal, and three intensities below optimal. Each stimulation intensity was systematically tested in three postures— supine (flat lying), semi-supine (45-degree inclined), and fully seated upright—in a randomized sequence. Each posture-intensity combination lasted five minutes, interspersed by a five-minute rest period to avoid carry-over effects.

Continuous physiological monitoring during the protocol included real-time transcutaneous carbon dioxide (CO_2_) measurements using Sentec Digital monitoring system (Timik Medical OY, Denmark) recorded at one-minute intervals, blood pressure monitoring, and room temperature control to minimize external confounders. After data collection completion, optical probes were carefully removed, and fNIRS data were subsequently analysed.

### fNIRS Measurement

The NIRSport2 fNIRS system used in this study provided scalability with 16 sources and 16 detectors allowing good coverage. Each unit offers high-powered dual-tip LEDs with up to 32mW illumination and advanced avalanche photodiode (APD) detectors, ensuring sensitivity down to 5 femtowatts. Signal integrity was further enhanced by proprietary automated optimization algorithms, short-distance detectors for extracerebral artifact control, and probe-level nine-axis accelerometers. The lightweight, compact design (approximately 900 grams, dimensions 162 mm × 125 mm × 60 mm) facilitated patient comfort and secure placement during measurements.

### fNIRS Data Analysis

Changes in oxyhemoglobin (HbO) and deoxyhemoglobin (HbR) concentrations were recorded using Aurora fNIRS Recording Software (NIRx Medical Technologies, LLC, Glen Head, NY, USA) and analysed using the *nirs*.*modules* in NIRS Brain AnalyzIR Toolbox (MATLAB-based, Mathworks Inc., USA)^15^.

Data Loading: Data imported using *nirs*.*io*.*loadDirectory*, structured by participant, group (three postures), and stimulation level.

Missing Data Handling: Corrected using *FixNaNs* for data continuity.

Stimulus Alignment: Aligned via the *ChangeStimulusInfo* module to accurately integrate geko™ stimulation events.

Demographic Integration: Participant characteristics incorporated using *AddDemographics*.

Filtering: Physiological noise mitigated by band-pass filtering (0.01–2.0 Hz).

Artifact Correction: Motion artifacts minimized using principal component analysis (*PCAFilter*, parameter 0.8) and temporal derivative distribution repair (*TDDR*).

Optical Density Conversion: Data transformed using the *OpticalDensity* function.

Haemoglobin Calculation: Applied Beer-Lambert Law (BeerLambertLaw) to derive HbO and HbR concentrations.

### Outcome Measure

The primary outcome measures are the beta (β) values derived from the General Linear Model (GLM) analysis (*nirs*.*modules*.*AR_IRLS*) applied to functional near-infrared spectroscopy (fNIRS) data. The *nirs*.*modules*.*AR_IRLS* module runs a GLM using an autoregressive iterative least squares algorithm for correcting motion and serially correlated error in fNIRS data^15^. The transcutaneous CO_2_ time-series was resampled and added as regressors (*AddAuxRegressors*) to the GLM model. The β-values from GLM quantified the relationship between experimental conditions (predictors) and the observed hemodynamic responses, specifically changes in HbO and HbR concentrations. By estimating β-values for each channel and condition, we assessed the magnitude and direction of hemodynamic changes associated with NMES. Positive β-values indicated an increase in haemoglobin concentration relative to the baseline, while negative β-values suggested a decrease. Our approach allowed for the identification of brain regions significantly activated during NMES, while accounting for confounding effects of CO_2_, offering insights into the underlying neural mechanisms.

### Statistical Analysis

Individual-Level Analysis: Hemodynamic responses estimated using autoregressive iterative reweighted least squares (*nirs*.*modules*.*AR_IRLS*) with a 30-second pre-stimulus baseline window^15^.

Group-Level Analysis: Mixed-effects regression (*nirs*.*modules*.*MixedEffects*) employed, controlling for fixed effects (experimental conditions) and random effects (subject variability) with robust fitting. Outlier detection and removal were performed using the *RemoveOutlierSubjects* module in the NIRS Brain AnalyzIR Toolbox^15^. This module identifies subjects whose statistical influence on the group-level model exceeds our user-defined threshold (cutoff = 0.05), based on measures such as leverage and residual variance from the mixed-effects model. Subjects identified as outliers were excluded entirely (*allow_partial_removal* = false) to maintain consistency across contrasts and ensure robust group-level inferences. This approach minimizes the influence of atypical data while preserving the integrity of the overall analysis. To examine the influence of experimental factors on NMES-evoked hemodynamic responses, a multiple linear regression model was conducted with HbO beta values as the dependent variable^16^. The predictors included Affected Side (left, right, bilateral), Lesion Location (cortical vs subcortical), Size of the Largest Lesion (Small lesion size: <1.5cm, medium lesion size: 1.5cm-5cm, large lesion size: >5cm), Body Position (laying flat, semisupine, and sitting), Stimulation Level (one intensity level above optimal: *VTE optimal+1, VTE* optimal, *and* three intensities below optimal: *VTE optimal-1, VTE optimal-2, VTE optimal-3*), and Hemisphere (left vs. right).

Region of Interest (ROI) Analysis: Measurement channels grouped into anatomically defined ROIs (left and right hemispheres), analysed using *roiAverage* function. To assess the impact of hemisphere and chromophore type on NMES-evoked hemodynamic responses, a two-way analysis of variance (ANOVA) was conducted with ROI (Left vs. Right hemisphere) and haemoglobin type (HbO vs. HbR) as fixed factors, and beta values from the GLM as the dependent variable.

Data Visualization: Topographical maps and statistical parametric maps (SPMs) created using the *printAll* function in the NIRS Brain AnalyzIR Toolbox^15^, with significance determined by a false discovery rate (q<0.05).

### Safety Monitoring

Safety parameters were rigorously monitored throughout the trial, with detailed reporting mechanisms established for adverse events (AEs), serious adverse events (SAEs), and adverse device effects (ADEs). Systemic hypotension and any device-related complications were also systematically recorded and managed according to standard clinical protocols.

### Ethical Approval

Ethical approval for the study was obtained from the local Research Ethics Committee (REC reference number: 343056). All study procedures complied with the ethical standards set forth by the Declaration of Helsinki.

## Results

A total of 18 participants with the mean age of 76 years (SE:3 years; range: 37 years) were included in the study. The cohort featured both left- and right-hemisphere strokes with 10 (56%) males and 9 (44%) females. The average interval between stroke onset and measurement was 32 days (SE = 6 days; range 125 days), and stroke severity at the time of recording, as measured by NIHSS, ranged from 0 to 10 (mean = 4, SE = 1). Their baseline characteristics including brain scan findings and vascular imaging revealed in Table 1. This sample reflected a representative cross-section of older adults in the subacute phase of ischemic stroke recovery.

**Table 1:** Baseline Characteristics.

| History of Hypertension | 9 (50.0%) |
| --- | --- |
| History of Diabetes | 5 (27.8%) |
| History of Stroke/TIA | 3 (16.7%) |
| Known AF | 6 (33.3%) |
| History of Congestive Cardiac Failure | 2 (11.1%) |
| Current or Ex-smoker | 6 (33.3%) |
| OCSP Classification |  |
| TACS | 7 (38.9%) |
| PACS | 4 (22.2%) |
| LACS | 3 (16.7%) |
| POCS | 4 (22.2%) |
| Location of the main lesion |  |
| Cortical | 9 (50.0%) |
| Others | 9 (50.0%) |
| Size of the Infarct |  |
| >5 cm | 5 (28.0%) |
| 1.5 - 5 cm | 9 (50.0%) |
| <1.5 cm | 4 (22.0%) |
| Ipsilateral ICA Stenosis |  |
| <=50% | 16 (89.0%) |
| >50% | 2 (11.0%) |
| No. of ICAS |  |
| None | 4 (22.2%) |
| 1 | 11 (61.1%) |
| >1 | 3 (16.7%) |

As part of the quality assurance and statistical refinement process, 84 data points were identified and excluded as outliers using the *RemoveOutlierSubjects* module in the NIRS Brain AnalyzIR Toolbox as described in the Methodology section. These subjects were flagged due to their disproportionate influence on the mixed-effects model estimates, as defined by a cutoff threshold of p<0.05. The removed participants spanned across multiple groups (see supplementary materials), suggesting systematic deviations potentially due to artifacts or physiological heterogeneity. This targeted removal ensured robust group-level inferences by reducing the effect of statistical leverage from anomalous data patterns. The final analysis was conducted on the refined dataset with improved homogeneity and interpretability.

The ROI analysis revealed no significant main effect of ROI on beta values, F(1, 1188) = 1.14, p = 0.285, indicating that activation levels did not differ significantly between left and right hemispheric regions (data file RETRAINresultsAR-IRLS-Laterality.xls in Supplementary Materials). Similarly, there was no significant main effect of haemoglobin type, F(1, 1188) = 1.10, p = 0.294, suggesting that the hemodynamic responses to NMES were comparable between oxygenated and deoxygenated haemoglobin. Importantly, the interaction between ROI and haemoglobin type was also non-significant, F(1, 1188) = 0.20, p = 0.653, indicating that the influence of ROI on activation did not depend on the chromophore measured. The statistical analysis did not reveal significant main effects on HbO beta values for Affected Side (F(2, 588) = 1.50, p = 0.224), Lesion Location (F(1, 588) = 1.53, p = 0.216), Hemisphere (F(1, 588) = 0.15, p = 0.703), Body Position (F(1, 588) = 0.02, p = 0.902), or Stimulation Level (F(1, 588) = 0.05, p = 0.827). However, Size of the Largest Lesion demonstrated a marginally significant effect on HbO beta values (F(1, 588) = 3.75, p = 0.053), suggesting a potential association between lesion size and the magnitude of HbO activation (data file LHBO.xls and RHBO.xls in Supplementary Materials).

Figure 2 displays topographical statistical parametric maps showing NMES-evoked changes in oxyhemoglobin (hbo) and deoxyhemoglobin (hbr) concentrations across the cortex with significance determined by a false discovery rate (q<0.05). The group-level GLM results indicate distinct lateralized and bilateral cortical activation patterns, with regions of both increased and decreased HbO and HbR responses in association with NMES stimulation (stim_channel1). These maps demonstrate a spatial distribution of cortical hemodynamic reactivity to NMES across all postures and peripheral stimulation levels.

Figure 3a illustrates the mean changes in combined (all channels) brain tissue oxygen saturation 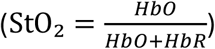 across three body positions (laying flat, semisupine, and sitting), under two experimental conditions: Baseline (no NMES) and Optimum NMES (individualized standard clinical setting for VTE). The x-axis represents body position, while the y-axis depicts the average (all channels) change in StO_2_. Positive values reflect an increase in oxygenation, whereas negative values indicate a decrease. Figure 3a bar graph data are presented as mean ± standard error. Comparisons between conditions (Baseline vs. Optimum) were performed within each posture. No significant differences in StO_2_ were observed between conditions in the laying flat (p = ns) and semisupine (p = ns) positions. However, in the sitting position, the Optimum condition resulted in a statistically significant increase in StO_2_ relative to Baseline (p = 0.010), indicating enhanced tissue oxygenation in this posture under the Optimum NMES condition. Also, linear modelling in the NIRS Brain AnalyzIR Toolbox revealed a modest positive trend in HbO beta values from supine to sitting posture (Figure S1 in supplementary materials). These results align with the physiological schematic presented in Figures 3b and 3c, suggesting NMES enhanced venous return and central hemodynamic in more upright positions, possibly facilitating improved brain tissue oxygen saturation from baseline.

**Figure 3:**
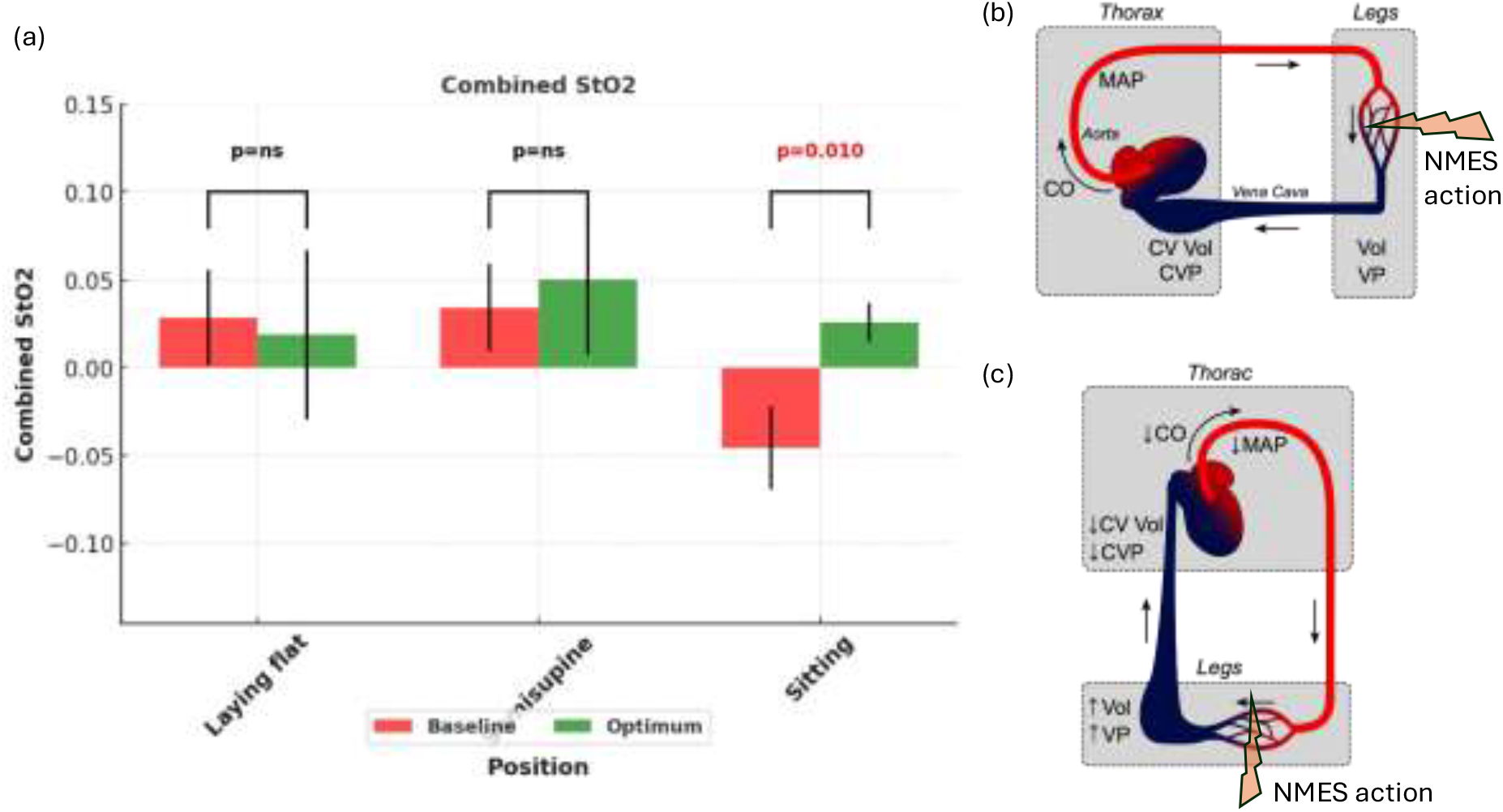
Posture related cardiovascular effects on cerebral tissue oxygenation (StO_2_) responses to neuromuscular electrical stimulation (NMES). (a) Bar graph showing changes in tissue oxygen saturation at three body positions (laying flat, semisupine, and sitting) under baseline (green) and optimum NMES (red) conditions. (b) Diagram of cardiovascular dynamics in the supine position. Venous return is maintained, supporting cardiac output (CO), mean arterial pressure (MAP), central venous pressure (CVP), and central venous volume (CV Vol), with lower venous pressure (VP) and volume (Vol) in the legs. (c) Diagram of cardiovascular changes in the upright posture, where gravitational pooling in the lower limbs reduces thoracic blood volume, leading to decreased CO, MAP, CVP, and CV Vol, and increased VP and Vol in the legs. (b) and (c) adapted from the original image from CVPhysiology.com that has been modified for illustrative purposes.

Figure S1 (supplementary materials) shows the effect of three body positions (laying flat, semisupine, and sitting) on the beta coefficients from GLM analysis of oxyhaemoglobin response. A linear mixed-effects model using the NIRS Brain AnalyzIR Toolbox shows how the beta values change across three body positions, controlling for other factors. There is a modest upward trend (red line), suggesting greater activation in the more upright posture (see Figure 3b and 3c for physiological interpretation).

Figure S2 (supplementary materials) shows the effect of the NMES levels (one intensity level above optimal: VTE optimal+1, VTE optimal, and three intensities below optimal: VTE optimal-1, VTE optimal-2, VTE optimal-3) on the beta coefficients from GLM analysis of oxyhemoglobin response. A linear mixed-effects model using the NIRS Brain AnalyzIR Toolbox shows how the beta values change across the NMES levels, controlling for other factors. There is an upward trend (red line), suggesting greater activation with higher NMES intensity. This finding supports a dose-dependent oxyhaemoglobin response, indicating that the intensity of NMES modulates the magnitude of cerebral oxygenation changes.

Figure S3 (supplementary materials) shows the effect of the size of the largest lesion (Small lesion size: <1.5cm, medium lesion size: 1.5cm-5cm, large lesion size: >5cm) on the beta coefficients from GLM analysis of oxyhaemoglobin response. A linear mixed-effects model using the NIRS Brain AnalyzIR Toolbox shows how the beta values change across the NMES levels, controlling for other factors. There is an upward trend (red line), suggesting greater activation with the increasing size of the largest lesion. A positive association whereby participants with large lesions (>5 cm) exhibited significantly greater HbO beta values. This may reflect impaired autoregulatory mechanisms or increased neurovascular demand in damaged brain regions. These findings align with prior literature indicating exaggerated hemodynamic responses in structurally compromised cortex during early stroke recovery^*17*^.

## Discussion

This study provides novel insights into the cerebral hemodynamic effects of transcutaneous NMES delivered via the geko™ device in subacute stroke survivors. Utilizing functional near-infrared spectroscopy (fNIRS), we observed significant increases in cortical oxygenation (HbO) during NMES, particularly in the upright seated posture. These findings suggest that NMES may enhance cerebral perfusion by augmenting venous return and cardiac preload^18^, thereby improving cerebral blood flow (CBF) in stroke patients.

A key observation was the significant increase in cortical tissue oxygenation during VTE optimal NMES, particularly in the upright seated posture. This finding suggests a synergistic effect between NMES-induced muscle pump activation and posture-related enhancement of venous return. In an upright sitting position, gravitational forces promote venous pooling in the lower extremities, reducing central venous volume and cardiac output; however, NMES appears to counteract this by enhancing venous return, potentially restoring cardiac preload and facilitating improved cerebral perfusion. These observations align with previous research indicating posture-dependent fluctuations in cerebrovascular parameters during stroke recovery and reinforce the role of the autonomic cardiovascular system in modulating CBF under physiological stressors^19^. The posture-dependent enhancement of cortical oxygenation aligns with other existing literature indicating that body position influences cerebral hemodynamic^20^ and this mechanism is supported by studies demonstrating that interventions increasing venous return positively impact cerebral perfusion in stroke patients^21^.

Our observation of a dose-dependent relationship between NMES intensity and HbO responses suggests that higher stimulation levels elicit greater cortical hemodynamic activation^11^. This finding supports our hypothesis that increased neuromuscular activation, reflected by augmented afferent input from the periphery, may trigger more robust neurovascular coupling responses in sensorimotor regions^11^. Here, the confounding effects of NMES-induced hypercapnia^13^ were accounted for and excluded from the GLM analysis. While the exact mechanisms underlying dose-dependent relationship between NMES intensity and HbO responses remain to be elucidated, it is also plausible that NMES enhances somatosensory integration and modulates local neuronal excitability, thereby influencing cerebral perfusion indirectly. Previous research has shown that peripheral stimulation can engage widespread cortical and subcortical circuits, potentially enhancing somatosensory integration and modulating local neuronal excitability, thereby influencing cerebral perfusion indirectly^22^,^23^.

Interestingly, participants with larger infarcts (>5 cm) exhibited greater HbO beta values in response to NMES. This could reflect impaired cerebral autoregulatory mechanisms or increased neurovascular demand in structurally compromised cortical areas. In such regions, NMES may elicit exaggerated hemodynamic responses due to diminished neurovascular reserve and compensatory increases in oxygen extraction. These results are congruent with fMRI and fNIRS studies reporting altered hemodynamic patterns in perilesional and contralesional cortices during early stroke recovery, particularly among individuals with extensive infarcts^24,25^.

The lateralized and bilateral activation patterns observed underscore the heterogeneity of stroke-related neural plasticity and perfusion dynamics^26^. Prior research has highlighted that post-stroke recovery is characterized by shifts in interhemispheric balance^27^, cortical excitability, and regional perfusion, which may influence functional neuroimaging^28^. The use of fNIRS in this study enabled sensitive detection of regional hemodynamic changes, offering a non-invasive and portable tool to monitor neurovascular responses in bedside environments. The integration of real-time CO_2_, temperature, and blood pressure monitoring further strengthened the reliability of hemodynamic interpretations by accounting for systemic confounders. Moreover, the rigorous statistical handling of artifacts and outliers using the NIRS Brain AnalyzIR Toolbox ensured the robustness of group-level inferences^15^.

Despite these strengths, several limitations warrant consideration. First, the modest sample size and the observational nature of this phase 1 study limit the generalizability of the findings. While the outlier removal process improved statistical fidelity, it may have inadvertently excluded physiologically relevant but heterogeneous responses. Future studies should employ larger cohorts with stratification by lesion characteristics, vascular status, and rehabilitation status to better delineate the factors influencing NMES responsiveness.

Second, the study did not include a sham or placebo stimulation condition, precluding the assessment of potential expectancy effects or non-specific activation. While the observed changes in HbO and HbR are consistent with physiologically plausible mechanisms, the inclusion of a sham-controlled arm in future trials would be essential for confirming the specificity of NMES effects on cerebral perfusion.

Third, although fNIRS offers excellent temporal resolution and is well-suited for monitoring superficial cortical activity, its spatial resolution is limited, particularly for deep structures such as the basal ganglia, thalamus, or brainstem. Combining fNIRS with complementary imaging modalities such as arterial spin labelling MRI or transcranial Doppler ultrasound could provide a more comprehensive picture of global and regional cerebral hemodynamics.

Fourth, while posture was varied, we did not account for individual differences in orthostatic tolerance or autonomic dysfunction, which are common after stroke and could influence hemodynamic stability. Incorporating measures of baroreflex sensitivity, cardiac output, or central venous pressure may enhance understanding of the cardiovascular-cerebral coupling under NMES.

Finally, although the study was conducted during routine clinical application of NMES for VTE prophylaxis, its extrapolation to neurorehabilitation requires further investigation. Specifically, future interventional studies should evaluate whether NMES-induced hemodynamic improvements translate into functional gains, such as motor recovery, gait performance, or cognitive outcomes. In this context, incorporating standardized clinical outcome measures alongside cerebral perfusion metrics will be crucial.

In conclusion, this study provides preliminary evidence that peripheral NMES via the geko™ device enhances cortical oxygenation in subacute stroke survivors, particularly in upright postures and at higher stimulation intensities. The hemodynamic responses appear modulated by lesion size and body position, indicating complex interactions between cardiovascular physiology and cerebral autoregulation post-stroke. These findings support the hypothesis that NMES may offer therapeutic benefits beyond VTE prevention, potentially serving as an adjunct to improve cerebral perfusion and promote recovery during the subacute phase of stroke rehabilitation^29^. In practical terms, implementing NMES early in the subacute phase may help maintain or improve CBF, particularly in patients unable to engage in active motor therapies due to significant weakness or fatigue. This is particularly relevant given that the early subacute period represents a critical window for neuroplastic reorganisation, where interventions that enhance cerebral perfusion and cortical engagement may be most impactful. Further controlled studies are warranted to confirm these effects and explore their functional implications^30^.

## Ethical Approval

The study was conducted in accordance with the Declaration of Helsinki and was approved by the local Research Ethics Committee (REC reference number: 343056) at the Stroke Rehabilitation Unit at Ellesmere Port Hospital, Chester, UK.

## Informed Consent

Written informed consent was obtained from all individual participants included in the study.

## Data Availability

The statistical results analysed during the current study are available in the supplementary materials.

## Acknowledgments

We would like to thank the patients and their families who generously participated in this study. We acknowledge the clinical and research staff for their invaluable support in recruitment and data collection. We are also grateful to the technical team at NIRx Medical Technologies for their support with the fNIRS equipment, and to Brain Products for assistance with EEG integration.

